# Does fixation of the right middle lobe/lingula during right upper lobectomy/left apical trisegmentectomy reduce rates of postoperative torsion/atelectasis?

**DOI:** 10.1101/2025.07.28.25332308

**Authors:** Devanish N. Kamtam, Douglas Z. Liou, Mark F. Berry, Satya Malla, Jake J. Kim, Natalie S. Lui, Irmina A. Elliott, Brandon Guenthart, Leah M. Backhus, Joseph B. Shrager

**Affiliations:** Division of Thoracic Surgery, Department of Cardiothoracic Surgery, Stanford University School of Medicine, CA, USA; Veterans Affairs Palo Alto Health Care System, Palo Alto, Stanford, CA, USA

**Keywords:** Middle lobe syndrome, Pulmonary atelectasis, Surgery, thoracoscopic, Robotic surgical procedures, Partial pneumonectomy

## Abstract

**Objective:** Frank torsion, or lesser atelectasis, of the right middle lobe/lingula may occur following right upper lobectomy/left apical trisegmentectomy. While some surgeons fix/pex the right middle lobe/lingula to the lower lobe, the necessity for this is unclear.

**Methods:** We retrospectively reviewed patients who underwent right upper lobectomy/left apical trisegmentectomy at our institution (2008-2023). We compared patients who underwent fixation vs. those who did not. The primary outcomes were incidence of acute torsion within 6 months following surgery and atelectasis on ∼6-month postoperative CT.

**Results:** Of the 497 patients, 438(88.1%) underwent right upper lobectomy and 143(28.8%) underwent fixation. Age, sex, race, comorbidities, and operative diagnosis were similar between the pexy and no-pexy groups (p=0.20-0.93). Thoracotomy approach was more common in the pexy group [64(44.8%) vs. 65(18.4%), p<0.001], whereas COPD [69(19.5%) vs. 10(7.0%), p<0.001) was more prevalent in the no-pexy group. On the primary outcomes, there was no difference between the groups in frank torsion [0(0%) vs. 0(0%)] or any atelectasis [23(22.5%) vs. 64(22.9%), p=0.94]. These findings were unchanged in subgroups of patients without COPD and with only right upper lobectomy patients. In the multivariate logistic regression model, undergoing fixation was not a significant predictor of atelectasis on 6-month postoperative CT (OR:0.94, 95% CI:0.48–1.86;p=0.87).

**Conclusions:** Fixation/pexy of the right middle lobe/lingula during right upper lobectomy/left apical trisegmentectomy does not substantially reduce rates of postoperative torsion or atelectasis. While assuring no malrotation of remaining lobes during lung re-expansion is likely important, the practice of pexy, which has downsides, should be largely abandoned.

**Central Picture:** 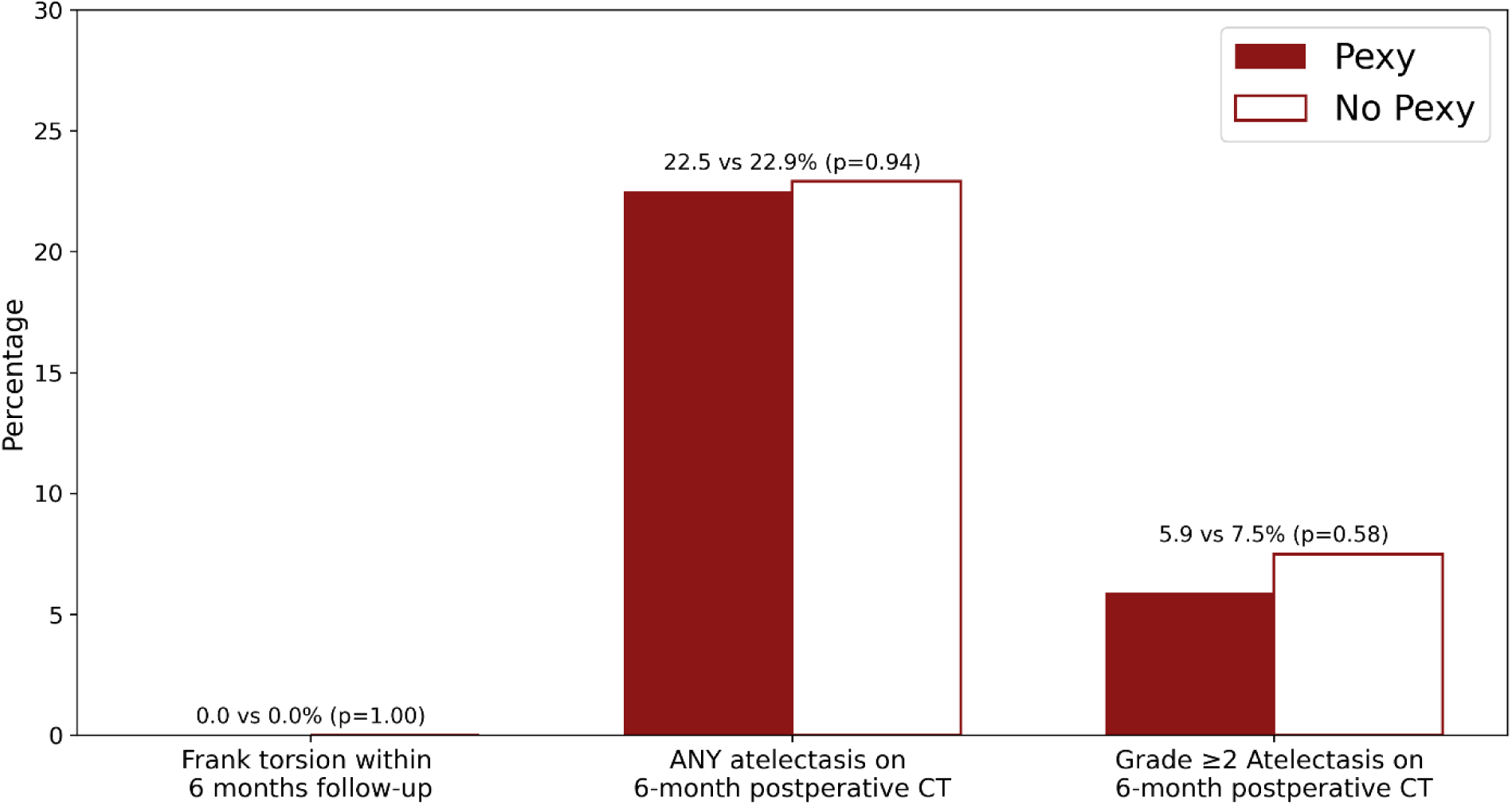

**Central picture legend:** No significant difference in frank torsion or atelectasis of middle lobe/lingula with pexy

**Central Message:** Routine prophylactic fixation of the right middle lobe/lingula to the lower lobe does not substantially reduce rates of postoperative torsion or atelectasis and may be abandoned as a routine practice.

**Perspective statement:** This large retrospective study evaluated the impact of prophylactic fixation (pexy) of the right middle lobe or lingula during “upper lobectomy.” Findings show no reduction in torsion, atelectasis, or early postoperative complications in the fixation group. Given the lack of benefit and potential risks, routine fixation should be reconsidered.

**Graphical abstract:** 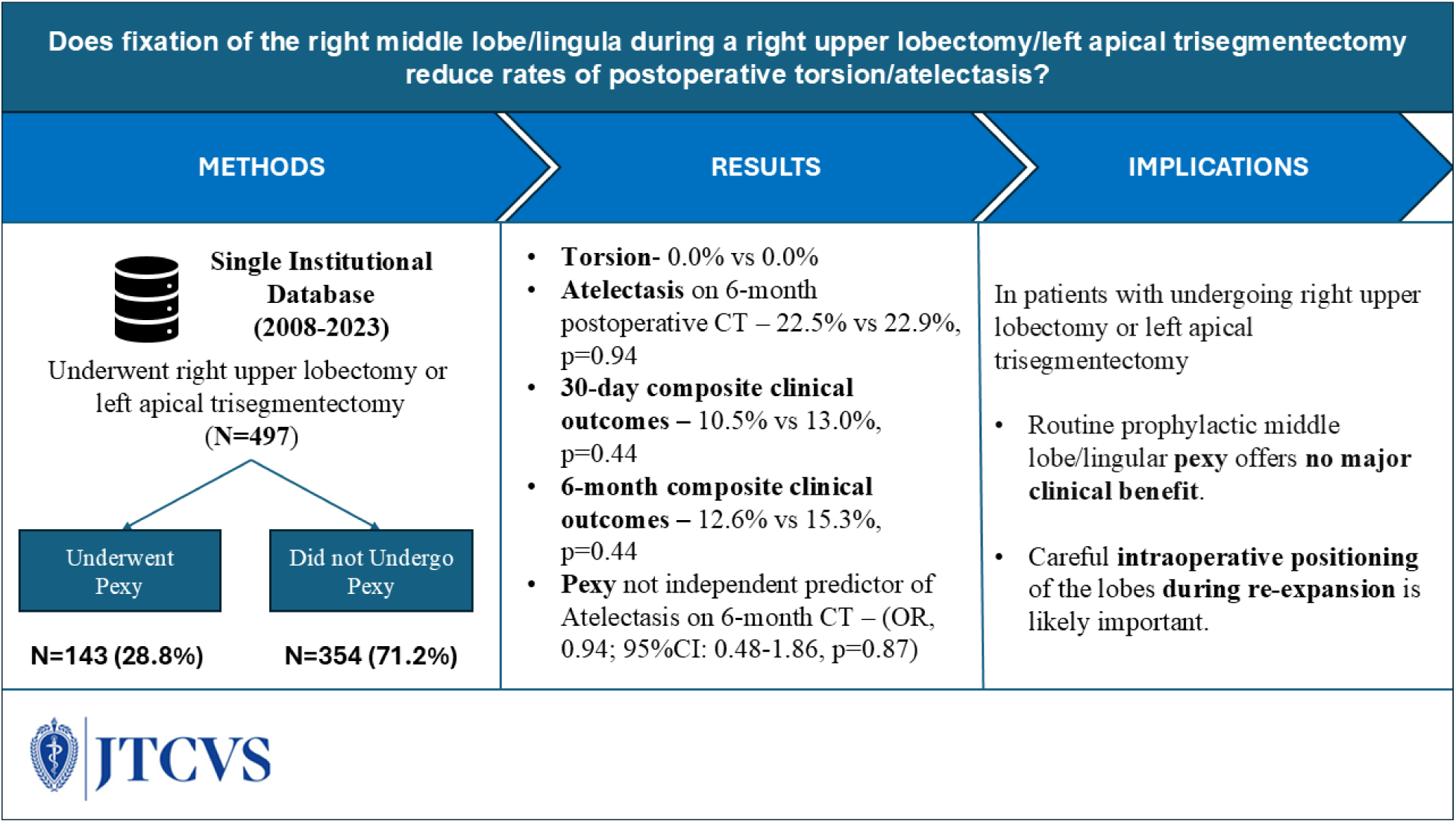

**Legend:** Graphical abstract summarizing this retrospective study evaluating the impact of prophylactic lobar fixation (pexy) during upper “lobectomy.”

## Introduction

According to the National Cancer Database, pulmonary lobectomies comprise approximately 70% of the 57,000 lung cancer resections performed annually in the USA^1^. Right upper and left upper lobectomies are the most commonly performed lung resections, comprising 31% and 23% of all lobectomies, respectively^2^. Left apical trisegmentectomy (LATS) is procedurally analogous to the right upper lobectomy (RULobectomy), as it preserves the lingula, which is anatomically similar to the right middle lobe. RULobectomy and LATS are uniquely associated with the rare yet potentially fatal complication of middle lobe (or lingular) torsion -- wherein a lobe axially rotates upon its bronchovascular pedicle.

Following a RULobectomy or LATS, the right middle lobe and lingula are susceptible to malrotation for several reasons. These include the small diameter and long length of their feeding bronchi, the frequent presence of a complete or near-complete major fissure allowing a great deal of mobility after division of the horizontal fissure (on the right) or the intersegmental plane (on the left), and a frequently sizeable residual apical space after the resection^3–6^. Lobar malrotation exceeding 180° can result in frank torsion, characterized by venous congestion, arterial compromise, and hemorrhagic infarction—a surgical emergency requiring immediate intervention. Lesser degrees of malrotation (<180°) may impair mucociliary clearance and lung expansion, leading to atelectasis-pneumonia, which may present with or without symptoms including chronic cough and/or dyspnea. This type of partial clockwise^7^ or anticlockwise^8,9^ rotation of the middle lobe following RULobectomy is one of the several appreciated causes of right middle lobe syndrome (RMLS). The overall incidence of frank lobar torsion following lobectomy has been reported at 0.08% to 0.3%^10^, with 70-85% of these cases involving RML torsion following RULobectomy^3,11^. Although frank torsion is rare, RMLS has been reported in approximately 8–11% of patients after RULobectomy^12,13^. The incidence of lingular torsion or lesser malrotation following LATS remains unknown^14–16^.

To mitigate the risk of frank torsion and lesser malrotation following RULobectomy/LATS, several prophylactic measures have been proposed and variably adopted by surgeons. These include careful positioning of the middle lobe during lung re-expansion, preservation of the inferior pulmonary ligament, and securing the middle lobe to the lower lobe using staplers, sutures, or adhesive strips^17–19—^ known as “pneumopexy” or simply “pexy”. Routine pexy has been proposed as an effective means to enhance stability of the RML/lingula and potentially reduce the risk of malrotation^9,10,13^. Others argue that careful lobar positioning during lung re-expansion is sufficient to mitigate this risk and that and that one should avoid unnecessary pleural trauma and potential air leaks^6,12,17,20,21^.

Outcomes data on the effectiveness of prophylactic pexy is limited to small case series with no control group^22^. Consequently, practice patterns vary widely—some surgeons advocate for routine pexy, others limit its use to patients with complete fissures or demonstrated excessive lobar mobility, and still others never pex.

This retrospective study utilized granular, single-institution data to determine whether pexy of the middle lobe or lingula reduces the incidence of frank torsion or lesser degrees of atelectasis following RULobectomy and LATS, assessed by their incidence within 6 months. Secondary objectives included evaluating the impact of pexy on broader postoperative morbidity and mortality.

## Methods

### Study design

This was a single-center, retrospective analysis approved by the Stanford School of Medicine Institutional Review Board (IRB), which waived the requirement for informed consent as the study involved only retrospective chart review (IRB-70048).

### Patient cohort

We retrospectively reviewed consecutive patients who underwent RULobectomy or left apical trisegmentectomy in the Division of Thoracic Surgery at Stanford University Hospital from January 2008 through December 2023. Patients were identified using the locally customized version of the Society of Thoracic Surgery (STS) General Thoracic Surgery Database, and those prior to September 2008 were identified from Stanford’s STARR clinical data warehouse. Data on demographics, comorbidities, operative diagnoses, surgical techniques (thoracotomy/video-assisted thoracoscopic surgery [VATS]/ robotic, pexy/no pexy), and histopathology findings were extracted from electronic medical records. Follow-up data were collected up to the standard, ∼6-month post-operative surveillance clinic visit.

Patients were categorized into two groups based on whether “pexy” of the right middle lobe or lingula to the lower lobe was performed (pexy group) or not (no-pexy group).

### Operative procedure

Operations were performed by 11 surgeons. Most were performed by VATS approach, with a smaller proportion utilizing robotic or open approaches (see Results). RULobectomy involved division of the horizontal fissure and posterior half of the major fissure, with maintenance of the anterior half of the major fissure. LATS involved division of the intersegmental plane between the lingula and the 3 apical segments and of the posterior half of the major fissure, with maintenance of the anterior half of the major fissure. In nearly all cases, division of the inferior pulmonary ligament was performed. When performed, “pexy” of the right middle lobe or lingula to the lower lobe utilized either a surgical stapler (in all minimally invasive and most thoracotomy cases) (Figure 1) or sutures (in a few thoracotomy cases) in an effort to prevent postoperative torsion or lesser malrotation. The decision to pex was made intraoperatively at surgeon’s discretion, based on surgeon habit, fissure completeness, perceived lobar mobility, and/or other unknown factors.

**Figure 1.**
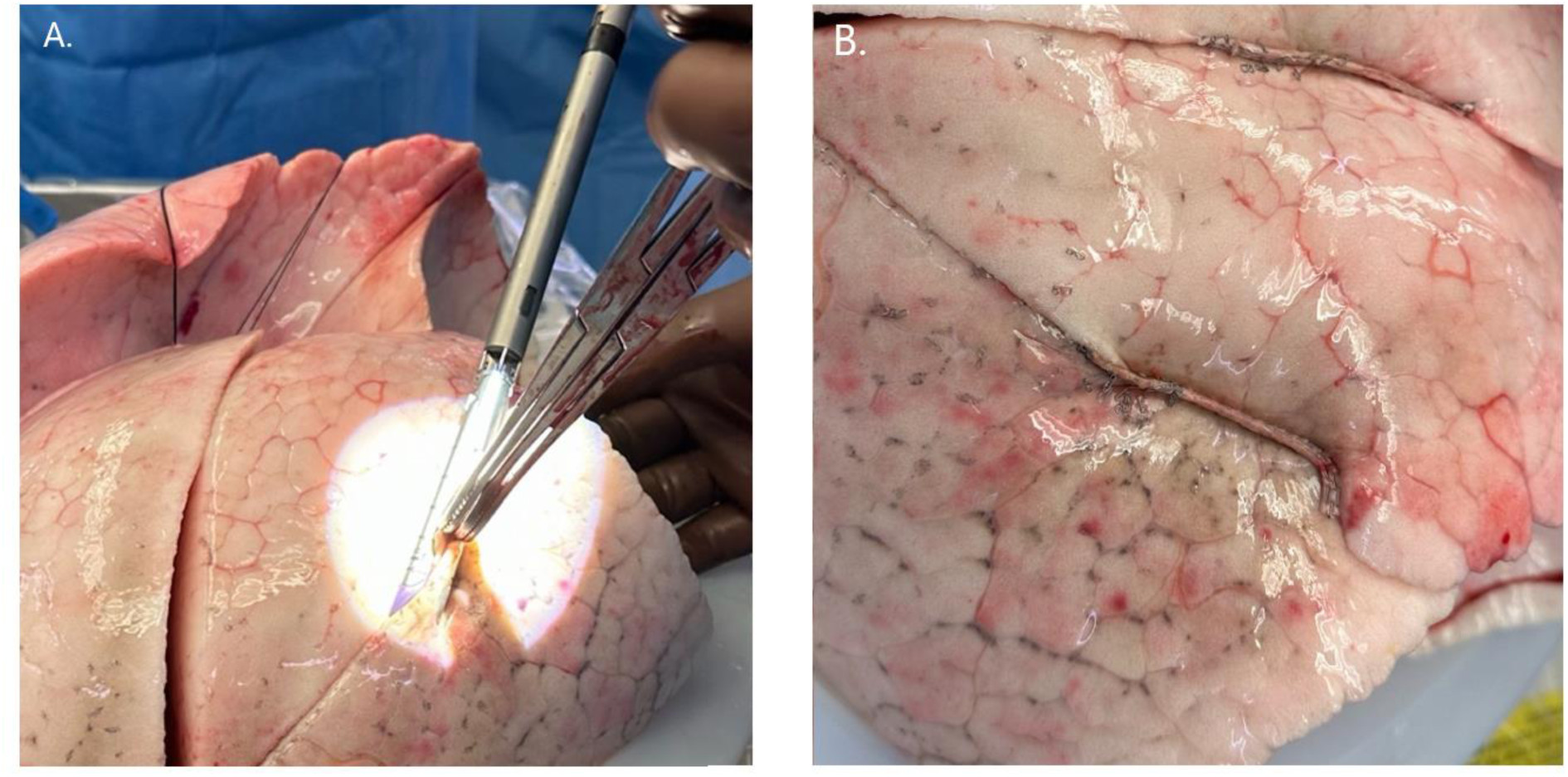
Representative images demonstrating pexy performed between lung lobes (not intraoperative images). **A.** Approximation and stapling of adjacent lung lobes using a clamp and Endo GIA stapler. **B.** Close-up view of stapled lobe-to-lobe pexy.

### Outcomes

Outcomes were assessed over a six-month postoperative period. The primary outcomes were the incidence of frank RML (or lingular) torsion within six months postoperatively, any RML (or lingular) atelectasis on the 6-month postoperative CT scan, and grade≥2 RML (lingular) atelectasis on 6-month postoperative CT scan, as determined by a review of CT scans by 2 surgeons. Frank torsion was defined as anatomic evidence of malrotation of the lobar bronchus with complete lobar atelectasis and substantial clinical deterioration.

Secondary outcomes included RML (or lingular) atelectasis on CXRs within 6 weeks postoperatively; a composite clinical outcome of mortality, prolonged length of stay (>14 days)^23^, unplanned readmission, reoperation, or bronchoscopy within 30 days and six months postoperatively; and the incidence of chronic cough at six months. Mortality, unplanned readmission, and reoperation were included only when attributable to pulmonary complications (e.g., acute respiratory distress syndrome, pneumothorax, hydro-pneumothorax, pleural effusion, subcutaneous emphysema).

### Radiological evaluation

CXRs obtained during the 6 postoperative weeks were classified as showing atelectasis when noted in the radiology report and described as being in the right upper lung field or of the right middle lobe.

RML (or lingular) atelectasis was assessed on the ∼6-month surveillance CT that all patients receive in our practice. Atelectasis was graded based on the extent of middle lobe/lingular parenchymal collapse observed on the scans upon review, and agreement, by 2 surgeons. A five-point scale was used: no atelectasis, minimal, sub-segmental, segmental, and lobar, modified from Newman et al^24^ (Figure 2).

**Figure 2.**
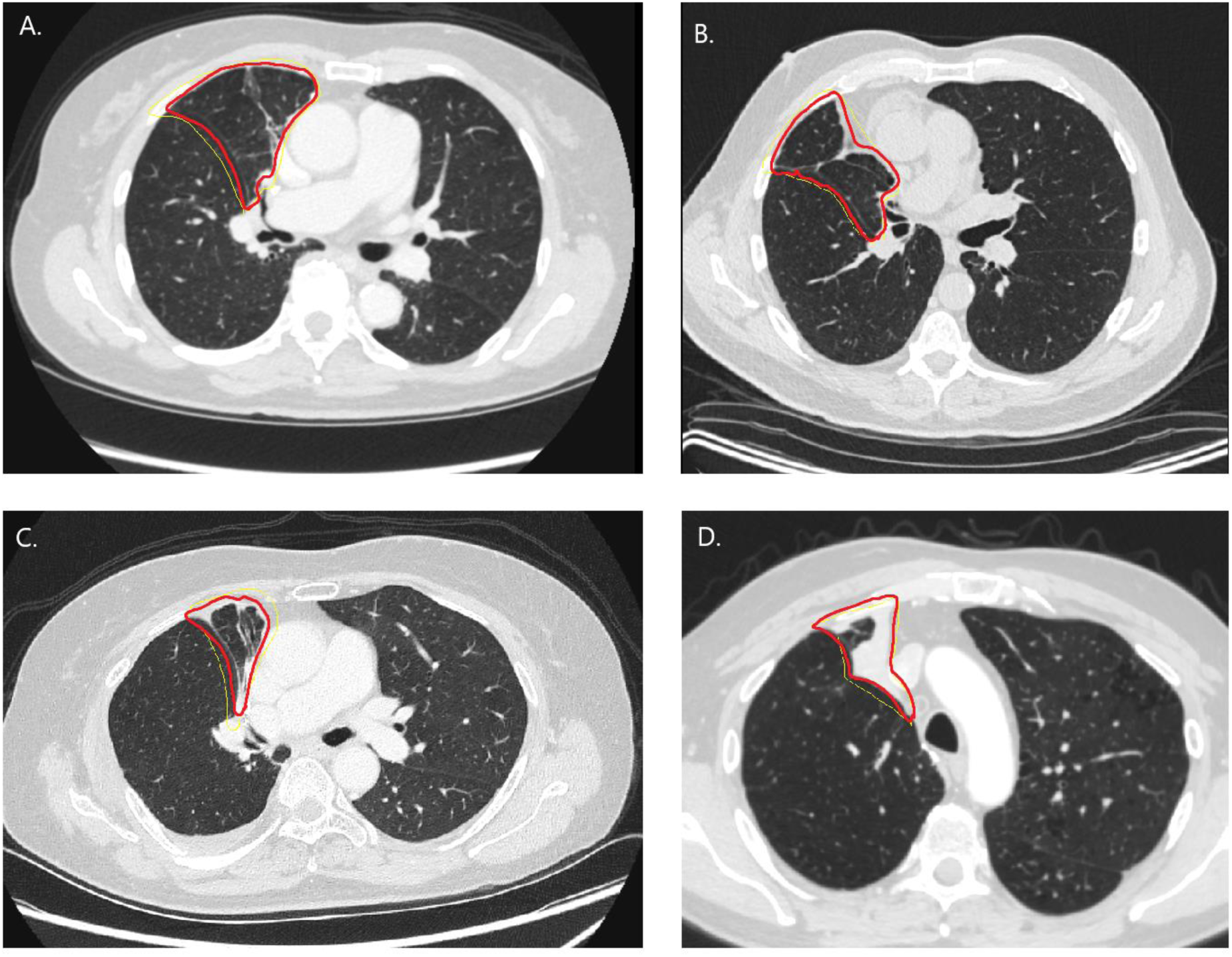
Axial CT images of 4 different patients obtained at ∼6 months postoperatively demonstrating the grades of atelectasis involving the right middle lobe, outlined in red. **A.** Minimal atelectasis—mild linear or platelike density with preserved aeration. **B.** Subsegmental atelectasis—wedge-shaped opacity involving a small subsegmental region. **C.** Segmental atelectasis— near-complete/complete collapse of a segment within the affected lobe. **D.** Lobar/sublobar atelectasis— near-complete/complete lobar collapse.

Fissure completeness was assessed based on the preoperative scan. The fissure between the RML (or lingula) and the respective lower lobe (i.e., the anterior half of the major fissure) was categorized as incomplete or complete based on axial CT sections with slice thickness ranging from 0.3 to 3mm. A fissure was classified as complete if it was distinctly visible as a sharp line for more than 95% of its extent.

All CT reviews were carried out blinded to clinical outcomes. RML atelectasis on postoperative CTs were validated by deep learning–based volumetry.

### Statistical analysis

Descriptive statistics were used to summarize baseline demographic, clinical, and radiological variables. The normality of continuous data was evaluated using the Shapiro-Wilk test. Normally distributed continuous variables were reported as mean ± standard deviation (SD) and compared using unpaired t-tests; non-normally distributed variables were reported as median (interquartile range, IQR) and compared using the Mann-Whitney U test. Categorical variables were presented as frequencies and percentages and compared using Pearson’s chi-square test (χ²) or Fisher’s exact test, as appropriate.

Univariate logistic regression was conducted to identify independent predictors of atelectasis on 6-month postoperative CT. Multivariable logistic regression models were developed using factors with p-value≤0.20 in univariate analyses, as well as “pexy” (regardless of its p-value).

Odds ratios (OR) with corresponding 95% confidence intervals (CI) were calculated. A two-tailed α<0.05 was considered statistically significant. All analyses were conducted using SPSS software (version 29.0, IBM Corp., Armonk, NY, USA).

## Results

### Baseline Cohort Characteristics

497 patients were included in the study: 438 (88.1%) underwent RUL and 59 (11.9%) underwent LATS. Pexy was performed in 143 patients overall (28.8%), including 136/438 (31.1%) of RULobectomies and 7/59 (11.9%) of LATS. Baseline patient demographics, comorbidities, surgical approach, and pathological characteristics stratified by pexy status are presented in Table 1. There were no significant differences in the distribution of age, sex, or race between the pexy and no-pexy groups. However, open surgical approach was significantly more common in the pexy group (44.8% vs. 18.4%, p<0.001). Conversely, COPD was significantly more prevalent in the no-pexy group (19.5% vs. 7.0%, p<0.001). Additionally, Charlson-comorbidity scores and the prevalence of liver disease were significantly higher in the no-pexy group (p=0.03).

**Table 1:** Baseline characteristics of patients stratified by whether or not they underwent pexy.

| Characteristics |  | Pexy |  | p-value |
| --- | --- | --- | --- | --- |
|  |  | Yes (n=143) | No (n=354) |  |
| Age (years) (Median, IQR) |  | 69.8(61.2-75.6) | 69.9(62.8-74.9) | 0.93 |
| Sex | Male | 58(40.6%) | 142(40.1%) | 0.93 |
|  | Female | 85(59.4%) | 212(59.9%) |  |
| Race | White | 91(63.6%) | 225(63.6%) | 0.41 |
|  | Black | 1(0.7%) | 7(2.0%) |  |
|  | Asian | 34(23.8%) | 83(23.4%) |  |
|  | Other | 12(8.4%) | 35(9.9%) |  |
|  | Unknown | 5(3.5%) | 4(1.1%) |  |
| Approach | Open | 64(44.8%) | 65(18.4%) | <0.001 |
|  | VATS | 78(54.5%) | 213(60.2%) |  |
|  | Robot-assisted | 1(0.7%) | 76(21.5%) |  |
| FEV<sub>1</sub> (%) (Median, IQR) |  | 94(80-108) | 97(78-108) | 0.93 |
| Diabetes mellitus |  | 17(11.9%) | 58(16.4%) | 0.20 |
| Chronic obstructive pulmonary disease |  | 10(7.0%) | 69(19.5%) | <0.001 |
| Congestive heart failure |  | 7(4.9%) | 21(5.9%) | 0.65 |
| History of cerebrovascular accident |  | 14(9.8%) | 36(10.2%) | 0.90 |
| Liver disease |  | 13(9.1%) | 58(16.4%) | 0.03 |
| Chronic kidney disease |  | 9(6.3%) | 40(11.3%) | 0.09 |
| Charlson-comorbidity score (Median, IQR) |  | 6(4-7) | 6(5-8) | **0.03** |
| Operative<br>Diagnosis | Adenocarcinoma | 104(72.7%) | 265(74.9%) | 0.80 |
|  | Squamous cell carcinoma | 16(11.2%) | 29(8.2%) |  |
|  | Neuroendocrine carcinoma | 5(3.5%) | 14(4.0%) |  |
|  | Mixed | 2(1.4%) | 10(2.8%) |  |
|  | Metastases | 7 (4.9%) | 18(5.1%) |  |
|  | Granulomatous inflammation | 4(2.8%) | 5(1.4%) |  |
|  | Other benign lesions | 5(3.5%) | 13(3.7%) |  |
| Complete fissures on CT |  | 23(26.4%) | 85(31.7%) | 0.35 |

FEV<sub>1</sub> – forced expiratory volume in 1 second; CT – computed tomography.

The overall incidence of frank lobar torsion was 0%. The overall incidence of any degree of atelectasis on CT at six months was 22.8% (87/381), with rates of 21.8% (72/331) following RULobectomy and 30.0% (15/50) following LATS. This atelectasis on CT was assessed at a median of 194 days (IQR, 180-208) following surgery.

### Comparison of Primary and Secondary Outcomes Between Pexy and No-Pexy Groups

Postoperative outcomes stratified by pexy status are presented in Table 2. There were no significant differences between the pexy and no-pexy groups in the incidence of torsion (0% vs. 0%) or atelectasis on six-month postoperative CT (22.5% vs. 22.9%, p=0.94). Additionally, the incidence of grade ≥2 atelectasis on 6-month postoperative CT was comparable between the two groups (5.9% vs. 7.5%, p=0.58), and the distribution of atelectasis grades showed no statistically significant difference (p = 0.51) (Figure 3). Almost all secondary outcomes, including composite clinical outcomes, prolonged length of hospital stay, mortality, readmissions, reoperations, and bronchoscopy within 30 days and 6 months postoperatively, as well as rate of chronic persistent cough at 6 months, were also not significantly different between the groups (Table 2, Figure 4). Only atelectasis on CXR within 6 weeks was significantly lower in the pexy group (21.1% vs 30.9% , p=0.03); but again, this did not correlate with a measurable difference in clinical outcomes .

**Figure 3:**
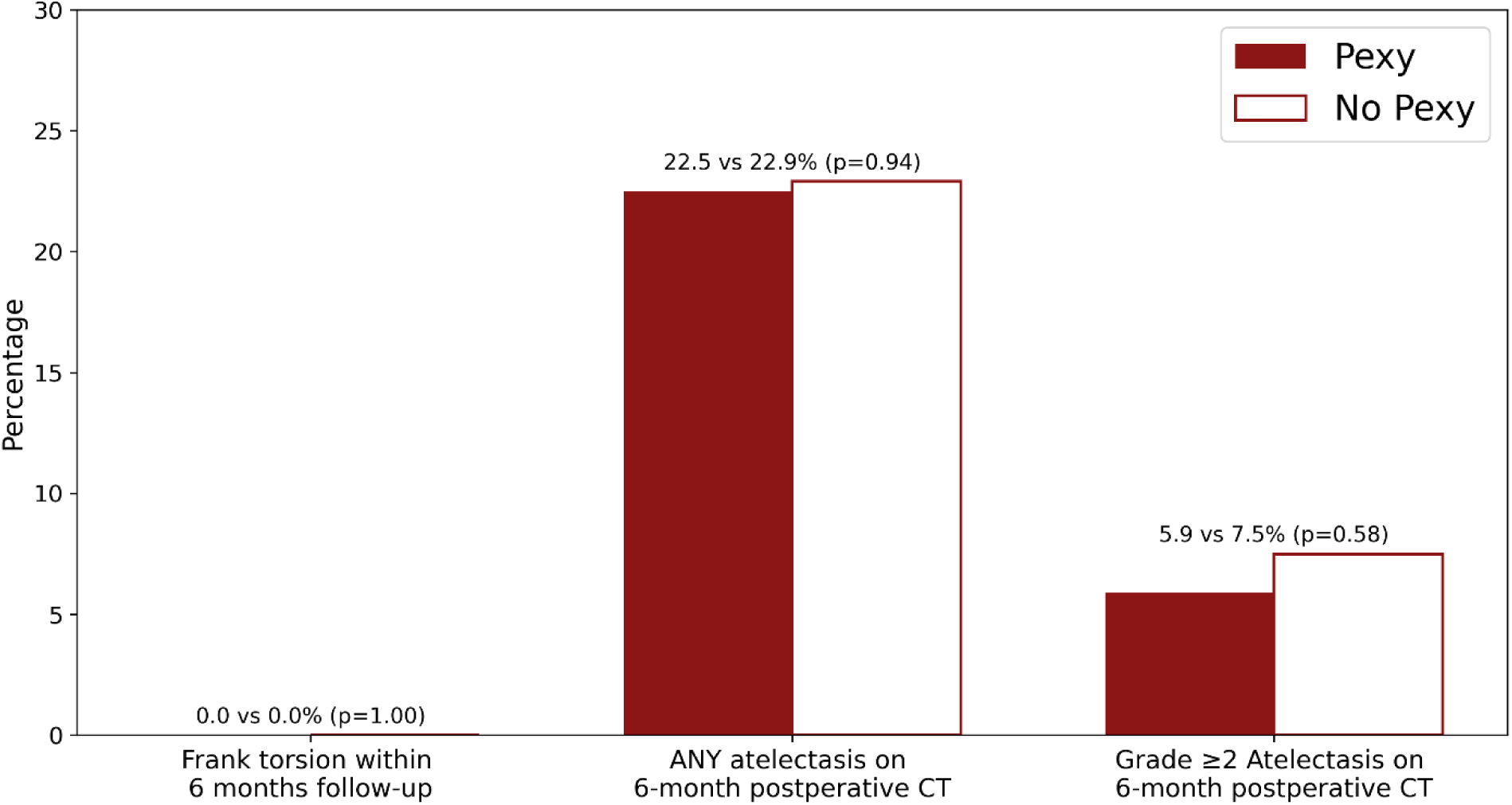
Primary outcomes of frank torsion within 6 months and atelectasis on 6-month follow-up chest CT in patients following right upper lobectomy or left apical trisegmentectomy. Note no significant differences in either outcome between the pexy and no-pexy groups.

**Figure 4:**
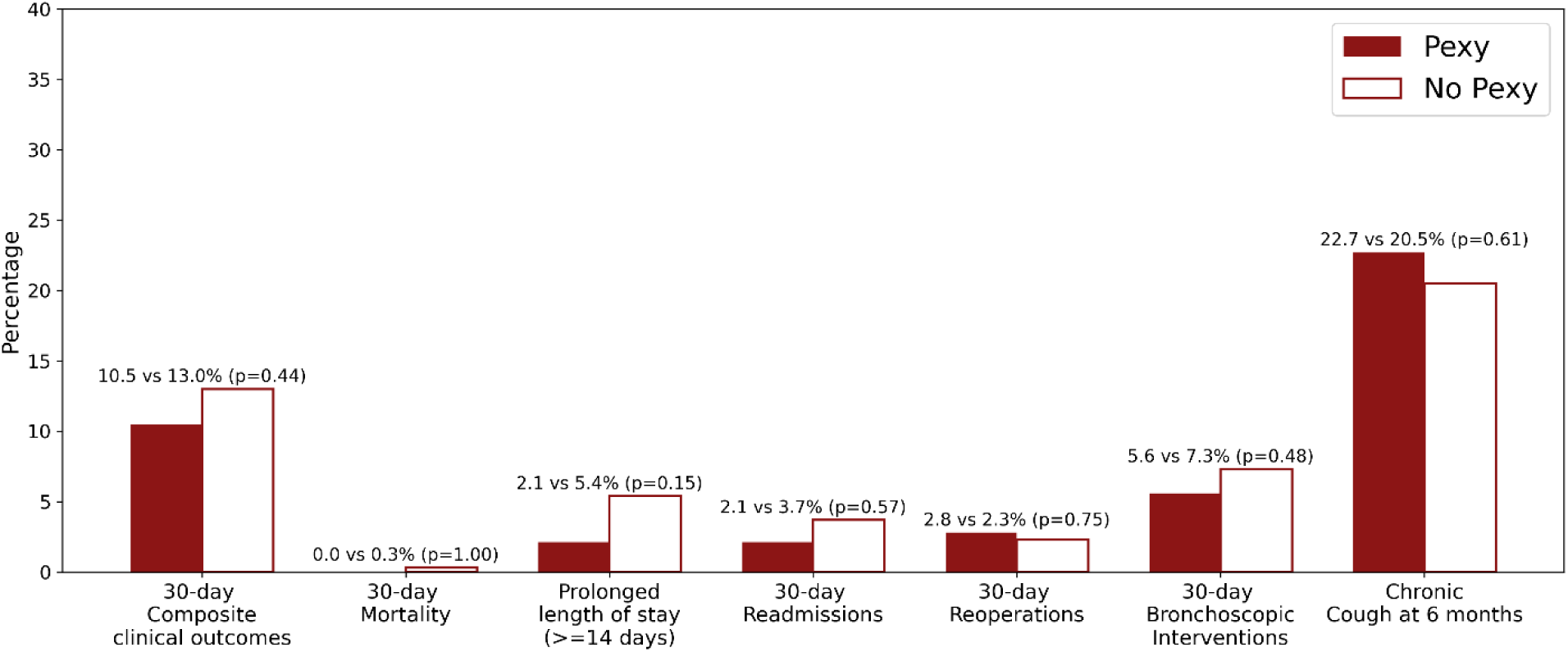
Secondary outcomes in patients who did or did not undergo pexy of the right middle lobe or lingula following right upper lobectomy or left apical trisegmentectomy, showing no significant differences across any measured outcomes.

**Table 2:**
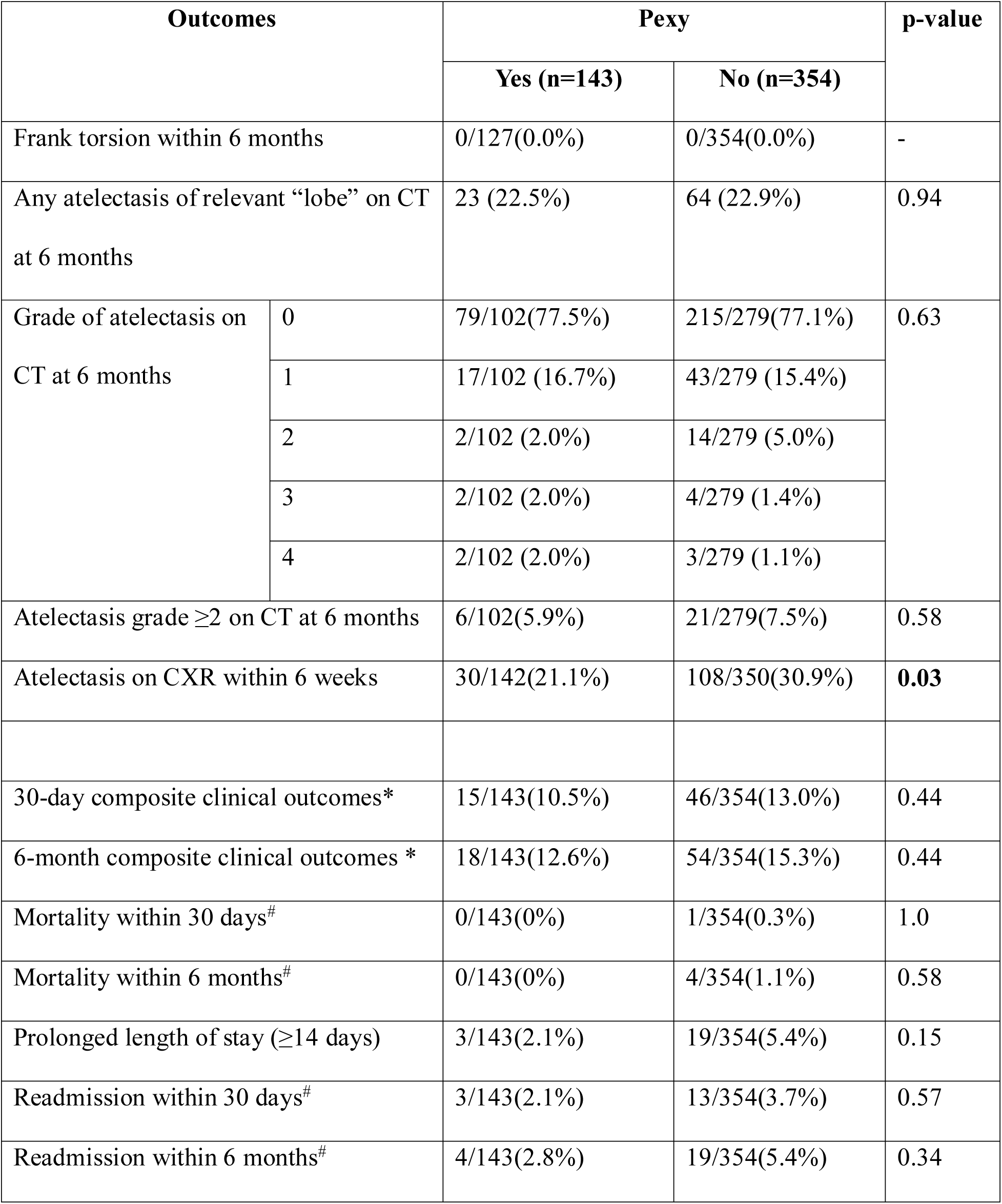
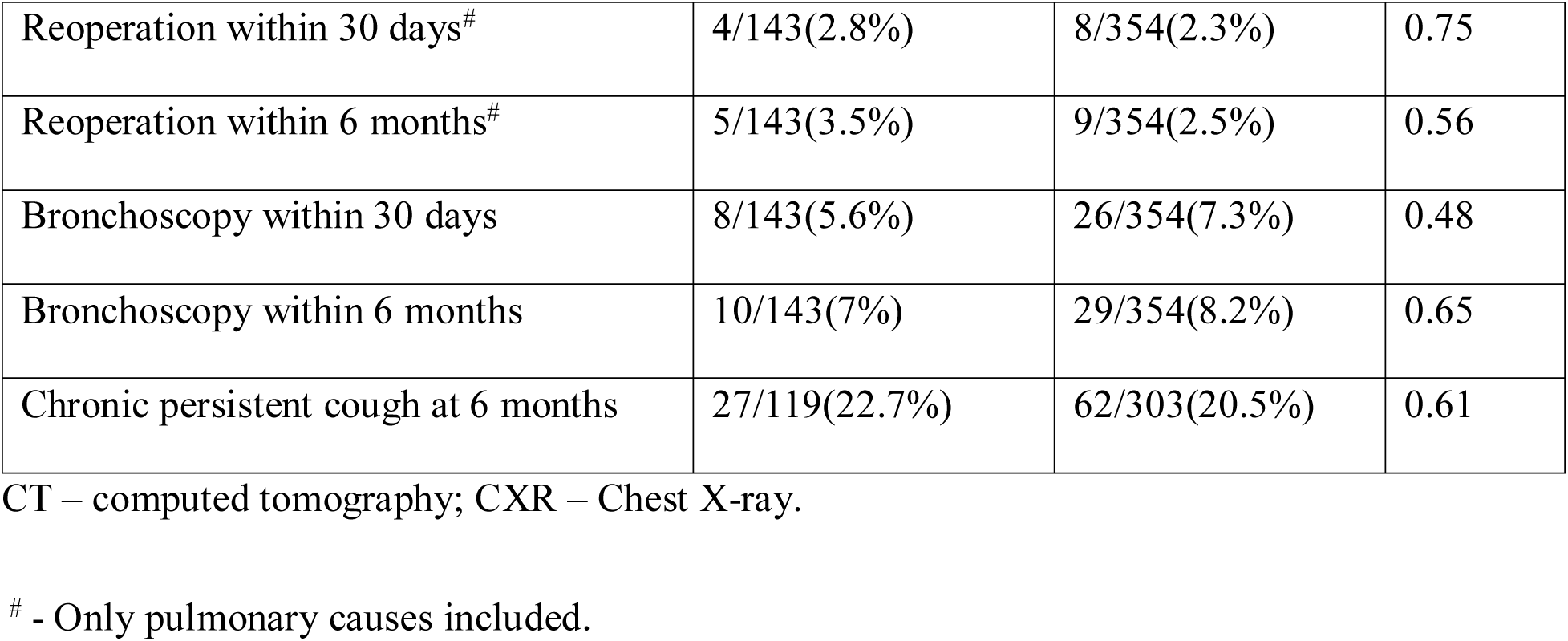
Postoperative outcomes in patients who underwent a right upper lobectomy or left apical trisegmentectomy stratified by whether or not they underwent pexy.

| Outcomes |  | Pexy |  | p-value |
| --- | --- | --- | --- | --- |
|  |  | Yes (n=143) | No (n=354) |  |
| Frank torsion within 6 months |  | 0/127(0.0%) | 0/354(0.0%) | - |
| Any atelectasis of relevant “lobe” on CT at 6 months |  | 23 (22.5%) | 64 (22.9%) | 0.94 |
| Grade of atelectasis on CT at 6 months | 0 | 79/102(77.5%) | 215/279(77.1%) | 0.63 |
|  | 1 | 17/102 (16.7%) | 43/279 (15.4%) |  |
|  | 2 | 2/102 (2.0%) | 14/279 (5.0%) |  |
|  | 3 | 2/102 (2.0%) | 4/279 (1.4%) |  |
|  | 4 | 2/102 (2.0%) | 3/279 (1.1%) |  |
| Atelectasis grade $\geq 2$ on CT at 6 months | | 6/102(5.9%) | 21/279(7.5%) | 0.58 |
| Atelectasis on CXR within 6 weeks |  | 30/142(21.1%) | 108/350(30.9%) | **0.03** |
| 30-day composite clinical outcomes* |  | 15/143(10.5%) | 46/354(13.0%) | 0.44 |
| 6-month composite clinical outcomes * |  | 18/143(12.6%) | 54/354(15.3%) | 0.44 |
| Mortality within 30 days<sup>#</sup> |  | 0/143(0%) | 1/354(0.3%) | 1.0 |
| Mortality within 6 months<sup>#</sup> |  | 0/143(0%) | 4/354(1.1%) | 0.58 |
| Prolonged length of stay ($\geq 14$ days) | | 3/143(2.1%) | 19/354(5.4%) | 0.15 |
| Readmission within 30 days<sup>#</sup> |  | 3/143(2.1%) | 13/354(3.7%) | 0.57 |
| Readmission within 6 months<sup>#</sup> |  | 4/143(2.8%) | 19/354(5.4%) | 0.34 |

|  |  |  |  |
| --- | --- | --- | --- |
| Reoperation within 30 days<sup>#</sup> | 4/143(2.8%) | 8/354(2.3%) | 0.75 |
| Reoperation within 6 months<sup>#</sup> | 5/143(3.5%) | 9/354(2.5%) | 0.56 |
| Bronchoscopy within 30 days | 8/143(5.6%) | 26/354(7.3%) | 0.48 |
| Bronchoscopy within 6 months | 10/143(7%) | 29/354(8.2%) | 0.65 |
| Chronic persistent cough at 6 months | 27/119(22.7%) | 62/303(20.5%) | 0.61 |

CT – computed tomography; CXR – Chest X-ray.
<sup>#</sup> - Only pulmonary causes included.

### Independent Predictors of Atelectasis on CT at 6-month postoperative follow-up

Univariate logistic regression analysis (Table 3) identified male gender (OR:0.71, 95%CI:0.44-1.15, p=0.17), history of congestive heart failure (OR:0.34, 95%CI:0.08-1.49, p=0.15), and complete fissures (OR:1.73, 95%CI:0.96-3.10, p=0.07) as potential predictors of atelectasis on six-month postoperative CT. After adjusting for all factors with p-values ≤0.20, none remained independent predictors. Performing a pexy was not a significant predictor (OR:0.94, 95%CI:0.48-1.86, p=0.87) of atelectasis on six-month postoperative CT.

**Table 3:**
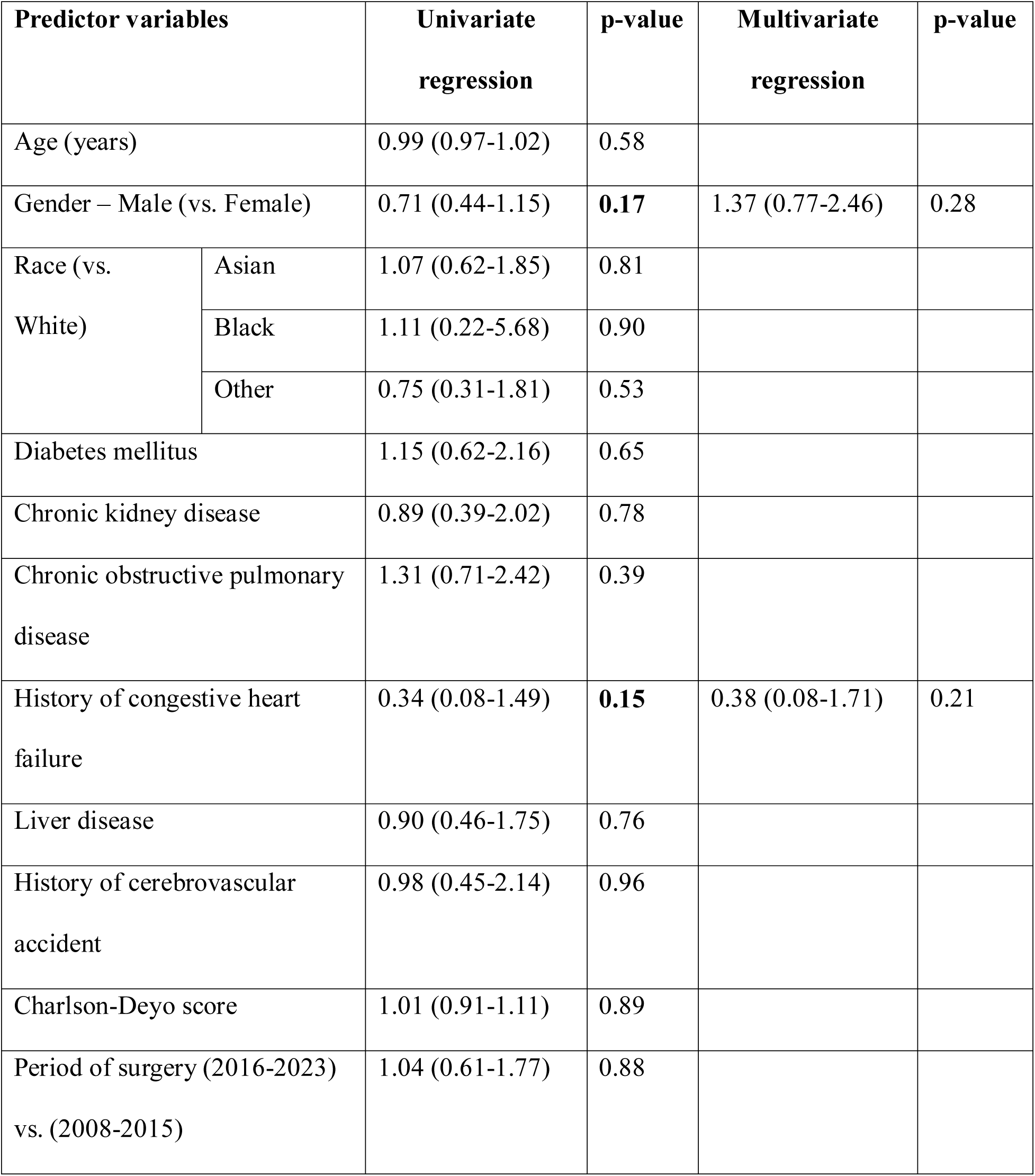
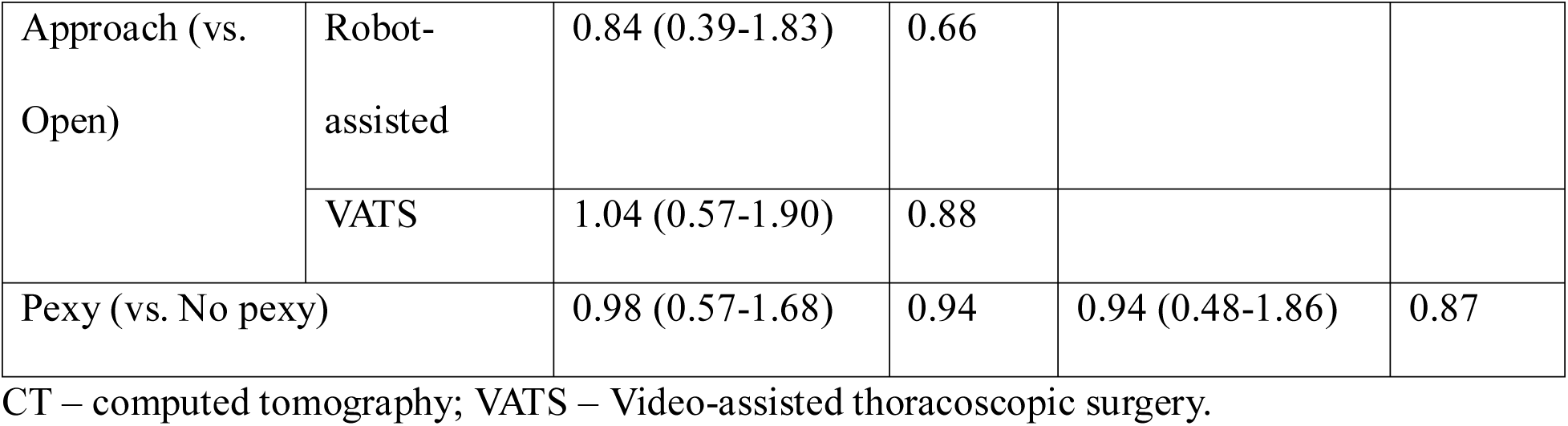
Logistic regression to determine independent risk factors of atelectasis on the ∼6-month postoperative CT following right upper lobectomy and left apical trisegmentectomy.

| **Predictor variables** |  | **Univariate**<br>**regression** | **p-value** | **Multivariate**<br>**regression** | **p-value** |
| --- | --- | --- | --- | --- | --- |
| Age (years) |  | 0.99 (0.97-1.02) | 0.58 |  |  |
| Gender – Male (vs. Female) |  | 0.71 (0.44-1.15) | **0.17** | 1.37 (0.77-2.46) | 0.28 |
| Race (vs.<br>White) | Asian | 1.07 (0.62-1.85) | 0.81 |  |  |
|  | Black | 1.11 (0.22-5.68) | 0.90 |  |  |
|  | Other | 0.75 (0.31-1.81) | 0.53 |  |  |
| Diabetes mellitus |  | 1.15 (0.62-2.16) | 0.65 |  |  |
| Chronic kidney disease |  | 0.89 (0.39-2.02) | 0.78 |  |  |
| Chronic obstructive pulmonary<br>disease |  | 1.31 (0.71-2.42) | 0.39 |  |  |
| History of congestive heart<br>failure |  | 0.34 (0.08-1.49) | **0.15** | 0.38 (0.08-1.71) | 0.21 |
| Liver disease |  | 0.90 (0.46-1.75) | 0.76 |  |  |
| History of cerebrovascular<br>accident |  | 0.98 (0.45-2.14) | 0.96 |  |  |
| Charlson-Deyo score |  | 1.01 (0.91-1.11) | 0.89 |  |  |
| Period of surgery (2016-2023)<br>vs. (2008-2015) |  | 1.04 (0.61-1.77) | 0.88 |  |  |
| Approach (vs. Open) | Robot-assisted | 0.84 (0.39-1.83) | 0.66 |  |  |
|  | VATS | 1.04 (0.57-1.90) | 0.88 |  |  |
| Pexy (vs. No pexy) |  | 0.98 (0.57-1.68) | 0.94 | 0.94 (0.48-1.86) | 0.87 |

CT – computed tomography; VATS – Video-assisted thoracoscopic surgery.

### Independent Predictors of 30-day composite clinical outcomes

Univariate logistic regression analysis (Supplementary Table 1) identified age (OR:1.06, 95%CI:1.03-1.09, p<0.001), race - Asian (OR:0.43, 95%CI:0.19-0.98, p=0.04), race – Black (OR:4.02, 95%CI: 0.93-17.48, p=0.06), Charlson-Deyo score (OR:1.12; 95%CI:1.01-1.24, p=0.02), surgical approach - robot-assisted (OR:0.56, 95%CI:0.25-1.25, p=0.15), and VATS (OR:0.34, 95%CI: 0.19-0.62, p<0.001), as potential predictors of 30-day composite clinical outcomes. After adjusting for all factors with p-values ≤0.20, age (OR:1.06, 95%CI:1.03-1.09, p<0.001) and VATS (OR:0.29; 95%CI:0.15-0.56, p<0.001) remained independent predictors. Performing a pexy was not a significant predictor (OR:0.66, 95%CI:0.32-1.34, p=0.25) of 30-day composite clinical outcomes.

### Subgroup Analyses

There was no significant difference in the incidence of torsion or atelectasis on six-month postoperative CT between the pexy and no-pexy groups, even among patients who underwent RULobectomy alone (excluding LATS). Similarly, nearly all other secondary outcomes within 30 days and 6 months postoperatively were also comparable between the two groups. Only atelectasis on CXR within 6 weeks was significantly lower in the pexy group (19.3% vs 29.8%, p=0.02) (Supplementary Table 2).

Among a subgroup of patients who underwent RULobectomy and had no preoperative diagnosis of COPD (which we thought might be a reasonable proxy for more complete fissures), there were no statistically significant differences between the pexy and no-pexy groups on any clinical or radiological outcome (Supplementary Table 3).

### CT volumetry Analyses

Deep learning-based volumetry revealed no significant difference between the pexy and no-pexy groups in the median reduction of normalized RML and right lower lobe (RLL) volumes between preoperative and postoperative scans, including RML/Right Lung volume (3.5% vs 2.8%, p = 0.47), RML/Total Lung volume (−0.4% vs 0.2%, p = 0.20), RLL/ Right Lung volume (33.6% vs 32.2%, p = 0.65), and RLL/ Total Lung volume (10.7% vs 11.9%, p = 0.31).

## Discussion

In this large, single-institution, retrospective cohort study, we found no significant difference in the incidence of frank lobar torsion or CT-graded atelectasis following RULobectomy/LATS at ∼ 6 months postoperatively between patients who underwent pexy and those who did not. This finding held true also in a subgroup who underwent only RULobectomy and one undergoing RULobectomy in the absence of COPD (thus perhaps with complete fissures). Other morbidity and mortality outcomes within 30 days and 6 months were also comparable between the pexy and no-pexy groups. A multivariable logistic regression analysis also did not identify having undergone pexy as an independent predictor of atelectasis.

These findings suggest that routine, prophylactic fixation of the middle lobe or lingula is not indicated to mitigate the risk of postoperative lobar malrotation to prevent either catastrophic frank torsion nor the lesser degrees of atelectasis that can result in annoying chronic cough or recurrent infections. Other patient-specific or surgical factors, rather than surgical fixation, appear to play a more significant role in the outcomes related to these postoperative complications. Studies evaluating prophylactic fixation techniques—particularly non-surgical approaches such as fibrin glue/sealant application—have suggested success in preventing postoperative lobar malrotation, though only in limited case series^17^. Comparative data assessing fixation techniques, particularly the efficacy of surgical pexy in preventing torsion and atelectasis, has been lacking.

Frank torsion is such a rare event (in fact 0% in our study) that we focused more carefully on lesser degrees of malrotation that may cause RMLS (or its left-sided equivalent) presenting with atelectasis and cough. Our findings indicate that pexy does not substantially reduce the incidence of these problems. We favor one factor as explaining the lack of a demonstrable effect of pexy: without doubt, our surgeons, in the absence of pexy, carry out careful positioning of the middle lobe/lingula, under direct visualization, during re-expansion of the lung following resection. This maneuver alone may be sufficient to prevent clinically relevant malrotation in nearly all patients.

It is also possible that bronchial kinking due to upward displacement of the lower lobe (particularly after division of the inferior pulmonary ligament) as demonstrated by Ueda et al, plays a larger role in atelectatic problems following RULobectomy than true malrotation on the middle lobe or lingular axis, which may be preventable by pexy. According to Ueda et al. up to 42% of patients experience bronchial kinking following RULobectomy and this was associated with higher rates of dyspnea and chronic cough (76% vs 21%, p<0.01)^25^. Although the observed atelectasis rate of 22% in our study exceeds the 17% incidence reported in another study^6^, this discrepancy may be due to the inclusion of subclinical (minimal) cases.

It must be noted that in our study, early postoperative CXRs did show a higher incidence of upper lung field/RML/lingular atelectasis in the pexy than the no-pexy group. However, these radiological-only findings were unassociated with significant differences in composite clinical outcomes between the groups, and they were also not reflected in atelectasis seen on CT scan at 6 months.

Given its doubtful benefits, we believe that routine prophylactic pexy should be abandoned, as it may prolong operative time by up to 10 minutes and predispose to air leaks. If it is not useful, it should be removed from routine use. However, there may well be certain subgroups of patients in which pexy is helpful. For example, there is a lower proportion of COPD in patients with RML atelectasis following RULobectomy (13.6% vs. 22.6%, p=0.16)^6^, which likely reflects the lower likelihood of a complete major fissure in patients with COPD. We interpret the significantly lower COPD prevalence in the pexy vs. the no-pexy group in our study (7.0% vs. 19.5%, p<0.001) to reflect surgeon appreciation of less middle lobe/lingular mobility in patients with COPD than in others. Based on our findings, ensuring optimal residual lobe positioning during lung re-expansion, before closure of incisions, is very likely sufficient to prevent axial malrotation in the vast majority of patients.

The key strengths of this study include its large sample size, the availability of granular single-institution data, blinded radiological re-evaluation to minimize observer bias, multiple independent reviewers for grading radiologic outcomes, and the incorporation of automated lung volume measurements, which enhance the reliability of the radiological outcomes. However, several limitations must be considered when interpreting these findings. Firstly, the single-center nature of this study may limit the generalizability of findings. Secondly, the retrospective design introduces potential selection bias and unknown confounders due to the absence of standardized preoperative and intraoperative criteria for decision-making regarding pexy. Further, the extended study period adds potential confounding related to changes in perioperative care over time despite the inclusion of study period in the regression model. Although it might be argued that the study may be underpowered to detect a true difference that might exist between the groups, the relatively large sample size and the consistent lack of significant difference across multiple outcomes—including on multivariate analysis—strengthen the validity of our findings. Of note, a post hoc sensitivity power analysis was conducted to determine the minimum detectable effect size for the outcome of CT-based atelectasis. Assuming a significance level of 0.05 (two-sided), 80% power, and our group sizes of 143(pexy) and 354(no-pexy), this analysis demonstrated that the study was powered to detect an absolute difference of >12.5% in any atelectasis between the groups. This in our opinion renders the study sufficiently powered to detect differences that would be considered clinically significant. Certainly, however, given all of the above limitations, a prospective multicenter study, stratified by fissure completeness, would be ideal to conclusively determine the role of pexy in various subgroups.

## Conclusion

This study provides strong support for the abandonment of routine prophylactic pexy of the middle lobe after RULobetoymy and of the lingula after LATS. These procedures do not substantially reduce the incidence of postoperative atelectasis or pulmonary morbidities. Given the lack of demonstrated benefits and the potential drawbacks of pexy, surgeons should prioritize meticulous intraoperative lobar positioning during re-expansion and utilize pexy only selectively.

## Glossary of Abbreviations

ARDS: Acute respiratory distress syndrome
COPD: Chronic obstructive pulmonary disease
CT: Computed tomography
CVA: Cerebrovascular accident
CXR: Chest X-ray
FEV_1_: Forced expiratory volume in 1 second
LATS: Left apical trisegmentectomy
NSCLC: non-small cell lung cancer
RML: Right middle lobectomy
RMLS: Right middle lobe syndrome
RL: Right lung
RLL: Right lower lobe
RUL: Right upper lobectomy
VATS: Video-assisted thoracoscopic surgery

## Data Availability

All data produced in the present study are available upon reasonable request to the authors.

---

**Supplementary Table 1:** Logistic regression to determine independent risk factors of 30-day composite clinical outcomes following right upper lobectomy and left apical trisegmentectomy.

| **Predictor variables** |  | **Univariate**<br>**regression** | **p-value** | **Multivariate**<br>**regression** | **p-value** |
| --- | --- | --- | --- | --- | --- |
| Age (years) |  | 1.06 (1.03-1.09) | **<0.001** | 1.06 (1.03-1.09) | **<0.001** |
| Gender – Male (vs. Female) |  | 1.40 (0.82-2.40) | 0.22 |  |  |
| Race (vs.<br>White) | Asian | 0.43 (0.19-0.98) | **0.04** | 0.53 (0.22-1.23) | 0.16 |
|  | Black | 4.02 (0.93-17.48) | **0.06** | 3.76 (0.81-17.43) | 0.09 |
|  | Other | 1.38 (0.60-3.15) | 0.45 | 1.40 (0.58-3.39) | 0.45 |
| Diabetes mellitus |  | 1.45 (0.73-2.88) | 0.29 |  |  |
| Chronic kidney disease |  | 1.45 (0.65-3.27) | 0.36 |  |  |
| Chronic obstructive pulmonary<br>disease |  | 1.04 (0.50-2.15) | 0.91 |  |  |
| History of congestive heart<br>failure |  | 1.60 (0.59-4.39) | 0.36 |  |  |
| Liver disease |  | 0.89 (0.41-1.97) | 0.78 |  |  |
| History of cerebrovascular<br>accident |  | 1.42 (0.63-3.18) | 0.40 |  |  |
| Charlson-Deyo score |  | 1.12 (1.01-1.24) | **0.02** | 1.04 (0.58-3.39) | 0.52 |
| Period of surgery (2016-2023)<br>vs. (2008-2015) |  | 1.11 (0.63-1.96) | 0.72 |  |  |
| Approach (vs. Open) | Robot-assisted | 0.56 (0.25-1.24) | **0.15** | 0.51 (0.21-1.25) | 0.14 |
|  | VATS | 0.34 (0.19-0.62) | **<0.001** | 0.29 (0.15-0.56) | **<0.001** |
| Complete fissure (vs. Incomplete) |  | 1.17 (0.61-2.23) | 0.64 |  |  |
| Pexy (vs. No pexy) |  | 0.78 (0.42-1.46) | 0.44 | 0.66 (0.32-1.34) | 0.25 |

CT – computed tomography; VATS – Video-assisted thoracoscopic surgery.

**Supplementary Table 2:**
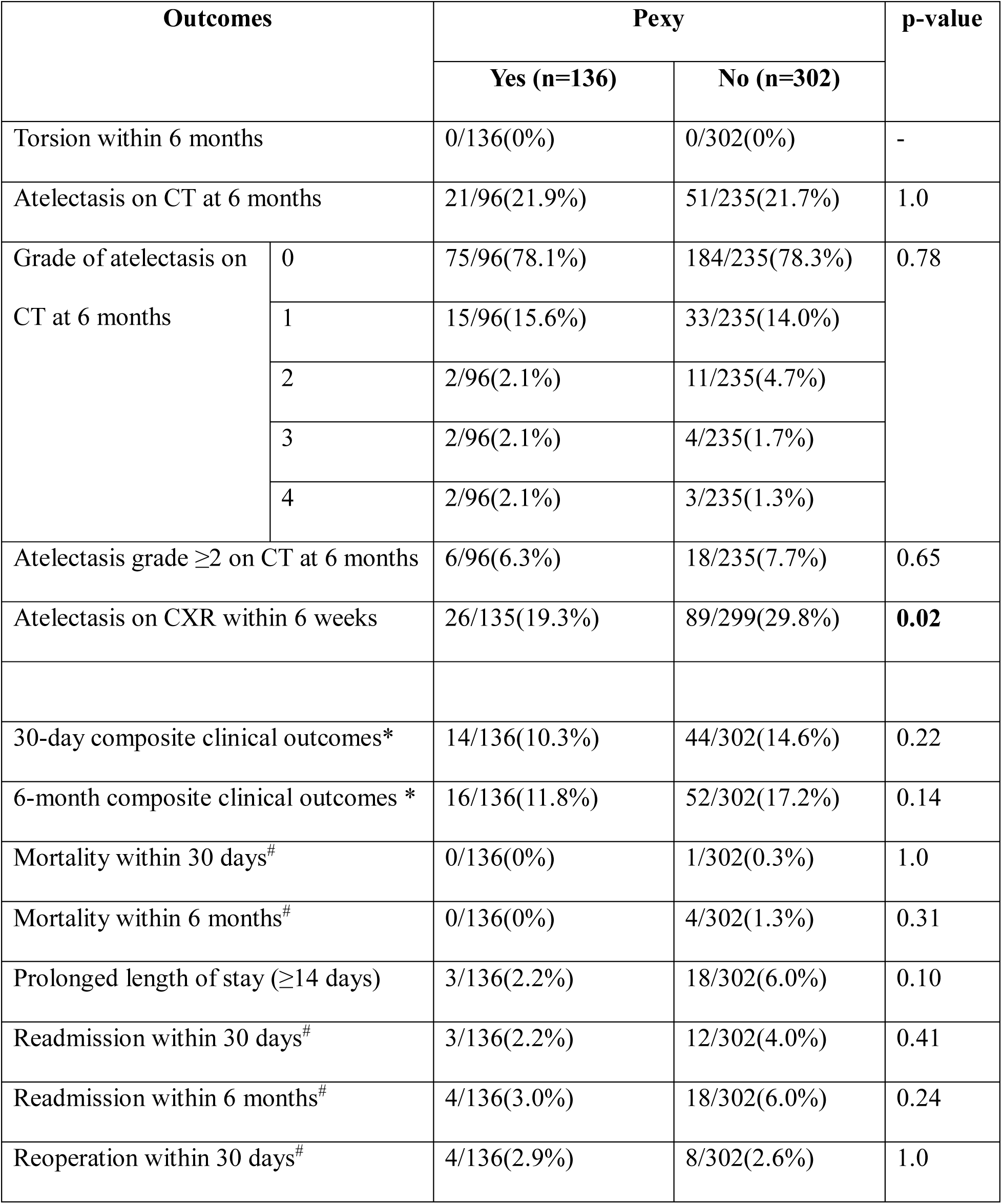
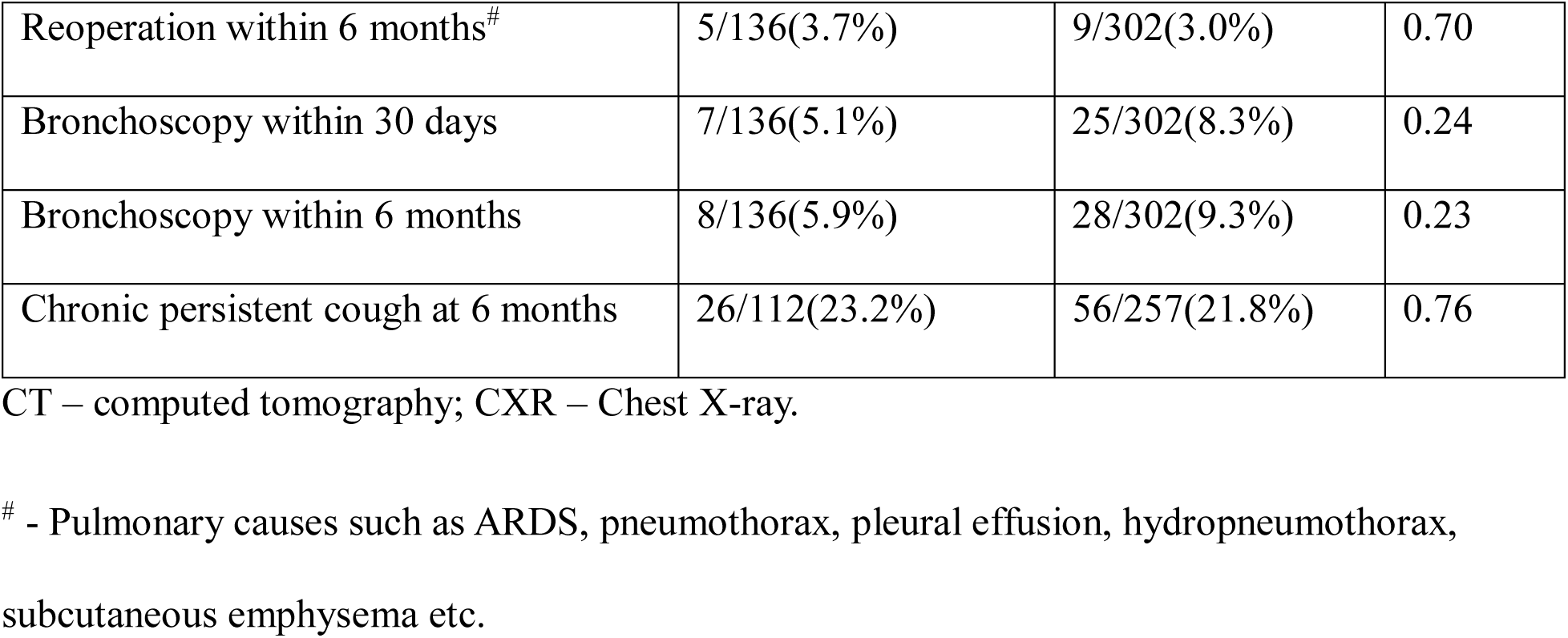
Postoperative outcomes in patients who underwent a right upper lobectomy (excluding left apical trisegmentectomies) stratified by whether they underwent pexy.

| Outcomes |  | Pexy |  | p-value |
| --- | --- | --- | --- | --- |
|  |  | Yes (n=136) | No (n=302) |  |
| Torsion within 6 months |  | 0/136(0%) | 0/302(0%) | - |
| Atelectasis on CT at 6 months |  | 21/96(21.9%) | 51/235(21.7%) | 1.0 |
| Grade of atelectasis on CT at 6 months | 0 | 75/96(78.1%) | 184/235(78.3%) | 0.78 |
|  | 1 | 15/96(15.6%) | 33/235(14.0%) |  |
|  | 2 | 2/96(2.1%) | 11/235(4.7%) |  |
|  | 3 | 2/96(2.1%) | 4/235(1.7%) |  |
|  | 4 | 2/96(2.1%) | 3/235(1.3%) |  |
| Atelectasis grade $\geq 2$ on CT at 6 months | | 6/96(6.3%) | 18/235(7.7%) | 0.65 |
| Atelectasis on CXR within 6 weeks |  | 26/135(19.3%) | 89/299(29.8%) | **0.02** |
| 30-day composite clinical outcomes* |  | 14/136(10.3%) | 44/302(14.6%) | 0.22 |
| 6-month composite clinical outcomes * |  | 16/136(11.8%) | 52/302(17.2%) | 0.14 |
| Mortality within 30 days<sup>#</sup> |  | 0/136(0%) | 1/302(0.3%) | 1.0 |
| Mortality within 6 months<sup>#</sup> |  | 0/136(0%) | 4/302(1.3%) | 0.31 |
| Prolonged length of stay ($\geq 14$ days) | | 3/136(2.2%) | 18/302(6.0%) | 0.10 |
| Readmission within 30 days<sup>#</sup> |  | 3/136(2.2%) | 12/302(4.0%) | 0.41 |
| Readmission within 6 months<sup>#</sup> |  | 4/136(3.0%) | 18/302(6.0%) | 0.24 |
| Reoperation within 30 days<sup>#</sup> |  | 4/136(2.9%) | 8/302(2.6%) | 1.0 |

|  |  |  |  |
| --- | --- | --- | --- |
| Reoperation within 6 months<sup>#</sup> | 5/136(3.7%) | 9/302(3.0%) | 0.70 |
| Bronchoscopy within 30 days | 7/136(5.1%) | 25/302(8.3%) | 0.24 |
| Bronchoscopy within 6 months | 8/136(5.9%) | 28/302(9.3%) | 0.23 |
| Chronic persistent cough at 6 months | 26/112(23.2%) | 56/257(21.8%) | 0.76 |

CT – computed tomography; CXR – Chest X-ray.
<sup>#</sup> - Pulmonary causes such as ARDS, pneumothorax, pleural effusion, hydropneumothorax, subcutaneous emphysema etc.

**Supplementary Table 3:**
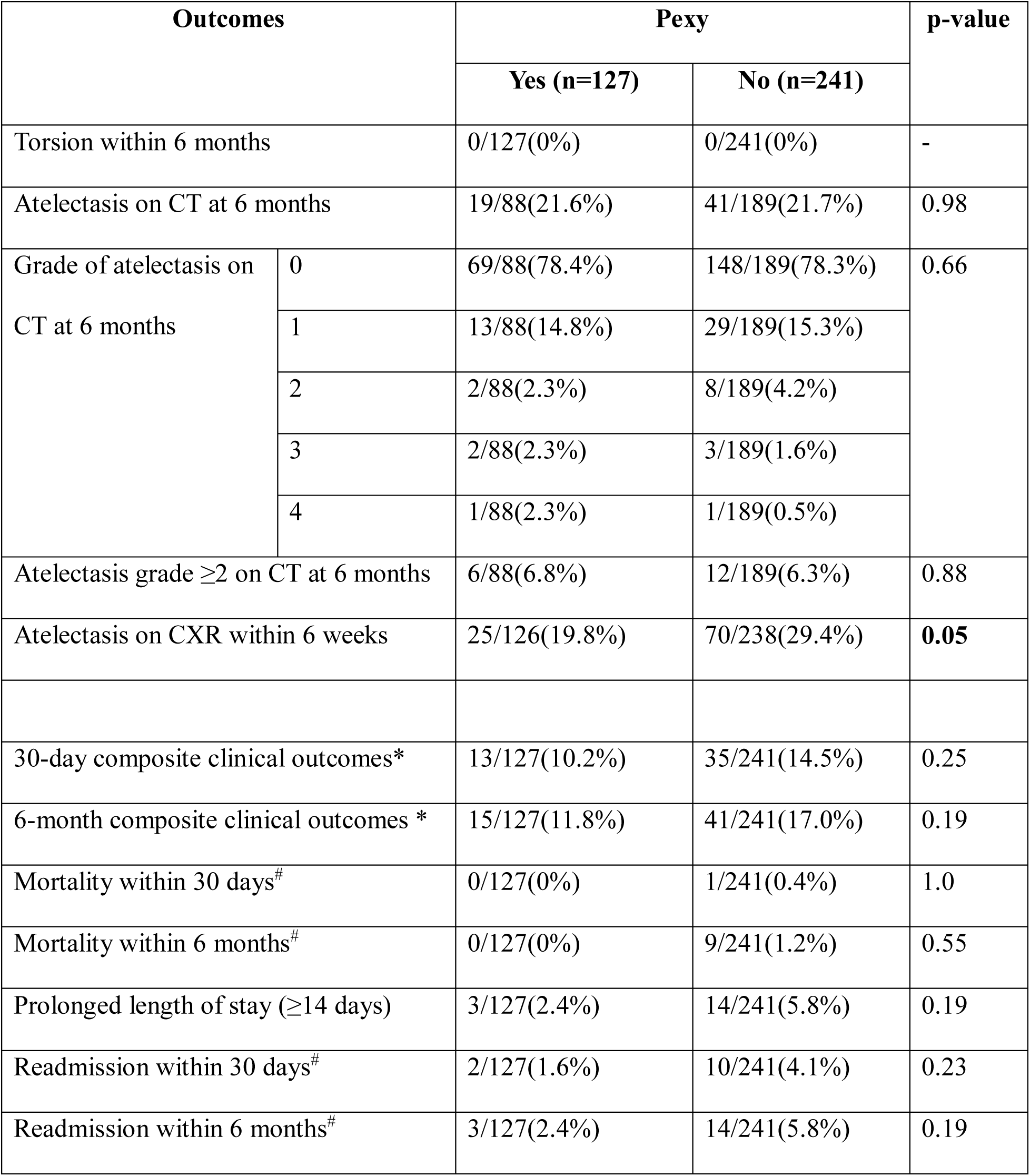
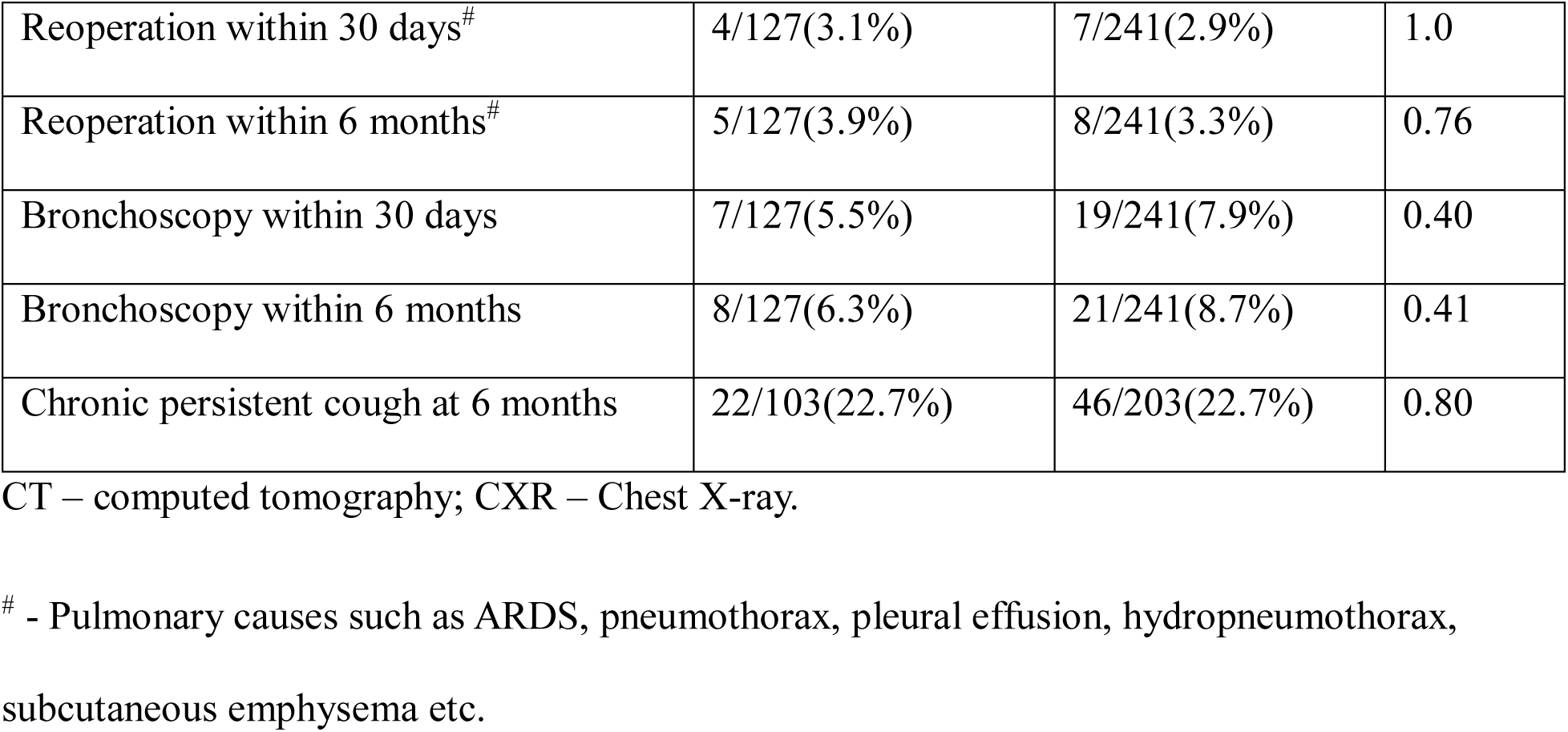
Postoperative outcomes in patients who underwent a right upper lobectomy and without a preoperative diagnosis of chronic obstructive pulmonary disease (COPD) stratified by whether they underwent pexy.

| Outcomes |  | Pexy |  | p-value |
| --- | --- | --- | --- | --- |
|  |  | Yes (n=127) | No (n=241) |  |
| Torsion within 6 months |  | 0/127(0%) | 0/241(0%) | - |
| Atelectasis on CT at 6 months |  | 19/88(21.6%) | 41/189(21.7%) | 0.98 |
| Grade of atelectasis on CT at 6 months | 0 | 69/88(78.4%) | 148/189(78.3%) | 0.66 |
|  | 1 | 13/88(14.8%) | 29/189(15.3%) |  |
|  | 2 | 2/88(2.3%) | 8/189(4.2%) |  |
|  | 3 | 2/88(2.3%) | 3/189(1.6%) |  |
|  | 4 | 1/88(2.3%) | 1/189(0.5%) |  |
| Atelectasis grade $\geq 2$ on CT at 6 months | | 6/88(6.8%) | 12/189(6.3%) | 0.88 |
| Atelectasis on CXR within 6 weeks |  | 25/126(19.8%) | 70/238(29.4%) | **0.05** |
| 30-day composite clinical outcomes* |  | 13/127(10.2%) | 35/241(14.5%) | 0.25 |
| 6-month composite clinical outcomes * |  | 15/127(11.8%) | 41/241(17.0%) | 0.19 |
| Mortality within 30 days<sup>#</sup> |  | 0/127(0%) | 1/241(0.4%) | 1.0 |
| Mortality within 6 months<sup>#</sup> |  | 0/127(0%) | 9/241(1.2%) | 0.55 |
| Prolonged length of stay ($\geq 14$ days) | | 3/127(2.4%) | 14/241(5.8%) | 0.19 |
| Readmission within 30 days<sup>#</sup> |  | 2/127(1.6%) | 10/241(4.1%) | 0.23 |
| Readmission within 6 months<sup>#</sup> |  | 3/127(2.4%) | 14/241(5.8%) | 0.19 |

|  |  |  |  |
| --- | --- | --- | --- |
| Reoperation within 30 days<sup>#</sup> | 4/127(3.1%) | 7/241(2.9%) | 1.0 |
| Reoperation within 6 months<sup>#</sup> | 5/127(3.9%) | 8/241(3.3%) | 0.76 |
| Bronchoscopy within 30 days | 7/127(5.5%) | 19/241(7.9%) | 0.40 |
| Bronchoscopy within 6 months | 8/127(6.3%) | 21/241(8.7%) | 0.41 |
| Chronic persistent cough at 6 months | 22/103(22.7%) | 46/203(22.7%) | 0.80 |

CT – computed tomography; CXR – Chest X-ray.
<sup>#</sup> - Pulmonary causes such as ARDS, pneumothorax, pleural effusion, hydropneumothorax, subcutaneous emphysema etc.

